# Identification of cutoff value for Homeostatic Model Assessment for insulin resistance (HOMA-IR index) in children at a tertiary care hospital of Bangladesh

**DOI:** 10.1101/2025.01.05.25320014

**Authors:** Ismat Jahan, Dhiraj Chandra Biswas, Farzana Sharmin, Roksana Parvin, Muhammad Rezaul karim, Suraiya begum

**Affiliations:** Upazilla Health Complex, Keraniganj, Dhaka, Bangladesh; Department of Paediatrics, Shahid Sayed Nazrul Islam Medical College, Kishorganj, Dhaka, Bangladesh; 100 Bedded District Hospital, Narsindi, Dhaka, Bangladesh; Department of Paediatrics, Khulna medical college hospital, Khulna, Bangladesh; OSD. Directorate General of Health Services, Dhaka; Department of Paediatrics, Bangabandhu Sheikh Mujib Medical University, Dhaka, Bangladesh

## Abstract

**Objective:** The Objective of this study was to find out the optimal cut-off value of HOMA-IR for identification of insulin resistance in Bangladeshi children.

**Material and Methods:** A cross sectional study was conducted at the department of Paediatrics of Bangabandhu Sheikh Mujib Medical University. 613 children aged 6-18years old were investigated for fasting insulin, glucose and lipid profile. HOMA-IR value was calculated from fasting insulin and fasting glucose level. The optimal cut-off value of IR was determined by percentile criteria and metabolic syndrome criteria.

**Results:** HOMA–IR cutoff value irrespective of age and sex at 85^th^ percentile ranges between 2.3 to 2.6 and cut off by metabolic syndrome criteria ranges between 2.0 to 2.4 with high sensitivity (93.2%) and specificity (60.1%) while HOMA-IR cut off at 95^th^percentile is higher (3.0 to 4.7) than the 85^th^centile but at 95^th^ centile sensitivity (78.6%) and specificity (72.9%) was low. Therefore suggested cut off value of HOMA-IR is 2.4 which is common in both metabolic syndrome criteria and percentile criteria.

**Conclusion:** This study suggests 2.4 as the optimal cut-off point of insulin resistance as because it is supported by both the percentile criteria and metabolic syndrome criteria.

## Introduction

Insulin resistance is a pathological condition characterized by lack of physiological response of peripheral tissues to insulin action (1,2). Insulin resistance and subsequent hyperinsulinemia is attributed to the development of serious health consequence such as obesity, type 2 diabetes mellitus, dyslipidemia, hypertension, atherosclerosis, polycystic ovarian syndrome, non-alcoholic fatty liver disease and cardiovascular disease (3). It has been argued that insulin resistance has a stronger relation to metabolic syndrome than even obesity and it is well established that insulin resistance is also present in patient without obesity(4).

IR may appear 1-2 decades before the formal diagnosis of type 2 diabetes and it was said that insulin resistance is the first sign of glucose intolerance (5,6) thus it plays a pivotal role in the pathogenesis of diabetes as well as metabolic syndrome.

There are different method used to detect insulin resistance among them the gold standard for the detection of IR is the euglycemic-hyperinsulinemic glucose clamp test (7), but as it is time consuming, expensive and invasive procedure and difficult to apply in larger scale epidemiological studies because it involves intra-venous infusion of insulin, taking frequent blood samples over a 3 hour period, and the continuous adjustment of a glucose infusion (8). So, alternative to the glucose clamp the most commonly used surrogate markers of insulin resistance is homeostatic model assessment method for insulin resistance (HOMA-IR index) which is simple, inexpensive and possible to study a large number of subjects with single glucose and insulin measurement in the fasting state (9).

Although the HOMA-IR has been widely used, it’s cut off values for insulin resistance differ for different ages, races, genders and diseases. Some authors used ROC curves for cut-off estimation (10–14) and some use percentile criteria (10,15–16). In our country optimal cut-off value of Insulin Resistance in children is absent. Therefore, the purpose of the study is to find out the optimal cut-off values of HOMA-IR for the diagnosis of IR in children.

## Materials and methods

### Study populations

This cross sectional study was conducted on 613 appearently healthy children between 6 to 18 years of age who visited Paediatric endocrinology clinic, Paediatric outpatient and inpatient department of Bangabandhu Sheikh Mujib Medical University from January 2019 to November 2020. Children taking systemic steroid for any cause, suffering from chronic disease like chronic kidney disease, chronic liver disease, pancreatic disease, endocrine disease (eg. Hypothyroidism, Cushing Syndrome, Hypothalamic disorder, Type 1 diabetes mellitus) and autoimmune disease, children use of medication (Sodium Valproate, Risperidon, Cyclosporine, Tarcolimus), hormonal replacement (Glucocorticoid, Growth hormone) which can interfere with glucose metabolism were excluded.

### Clinical and laboratory measurements

Weight and height was measured by using electronic weighing machine and stadiometer respectively to the nearest 0.1cm and 0.2kg. Waist Circumference (WC) was measured midway between the lowest rib and the superior border of the iliac crest by using a non-extensible and non-elastic measuring tape in mid respiration. BMI was calculated as weight (kg) divided by height squared (m2) and classified as normal weight if BMI was BMI ≥5th to ˂ 85^th^ percentile, over weight if BMI was 85 th to 95 th percentile, or obese if BMI was ˃95 th percentile according to CDC growth chart 2000 (19). Blood pressure was measured in the sitting position after at least 5 minutes of rest with appropriate size culf.

Pubertal status was assess according to Tanner stage, who were in tanner stage 1 are known as Pre- pubertal stage and who were in tanner stage 2, 3 and 4 were known as Pubertal stage (20,21)

Blood samples were obtained after an overnight fast of at least 8 hours, and biochemical assays including fasting insulin level, fasting blood glucose,2 hour after 1.75gm/kg glucose and fasting lipid profile were measured using Architect plus ci4100(Automated analyzer). Estimation of serum glucose by Glucose Oxidase (GOD-PAP) method, fasting serum insulin by Micro particle enzyme immunoassay (MEIA) technique, serum TG by Glycerol Phosphate Oxidase (GPO-PAP) method, serum HDL-C by Accelerator Selective Detergent method, serum total cholesterol by Cholesterol oxidase (CHOD-PAP) method, serum LDL-cholesterol by Frieldwald’s formula.

### Definition of Metabolic syndrome and calculation of HOMA-IR

In our study, we used modified ATP III criteria (Adult Treatment Panel III) in which MS was defined by presence of three or more of the following five components :Waist circumference≥ 90 th percentile for age and sex, HDL cholesterol 40 mg/dl or specific treatment for this lipid abnormality, Triglyceride ≥ 150 mg/dl or specific treatment for this lipid abnormality, Blood pressure ≥ 130/85 mm Hg or treatment of previously diagnosed hypertension, fasting plasma glucose ≥ 5.6 mmol/l or previously diagnosed type 2 diabetes (22).

HOMA-IR index was obtained by calculating the fasting insulin(microU/ml) and fasting blood glucose (mmol/L) divided by 22.5 (23).

### Statistical analysis

Data analysis was done by Statistical Program for Social Science software (SPSS, version 25.0). Data were presented in tables, figure and diagrams. Sensitivity, specificity, positive and negative predictive value were calculated at different cut-off points of the HOMA-IR in relation to metabolic syndrome by receiver operating chacteristic curve (ROC) curve. The Youden index (max of sensitivity+specificity) was calculated to identify the cutt-off point. Without metabolic syndrome the cut-off values for insulin resistance was based on percentile criteria.

### Ethics statement

Ethical clearance were taken from the Institutional Review Board (IRB), Bangabandhu Sheikh Mujib Medical University, Dhaka.

## RESULTS

In Table 1 Baseline characteristics was assessed and summarizes anthropometric, clinical, and biochemical characteristics of the study population. We included total 613 children in this study, among them 167 normal weight,171 over weight,275 Obese which shown in fig 1.

**Fig 1:**
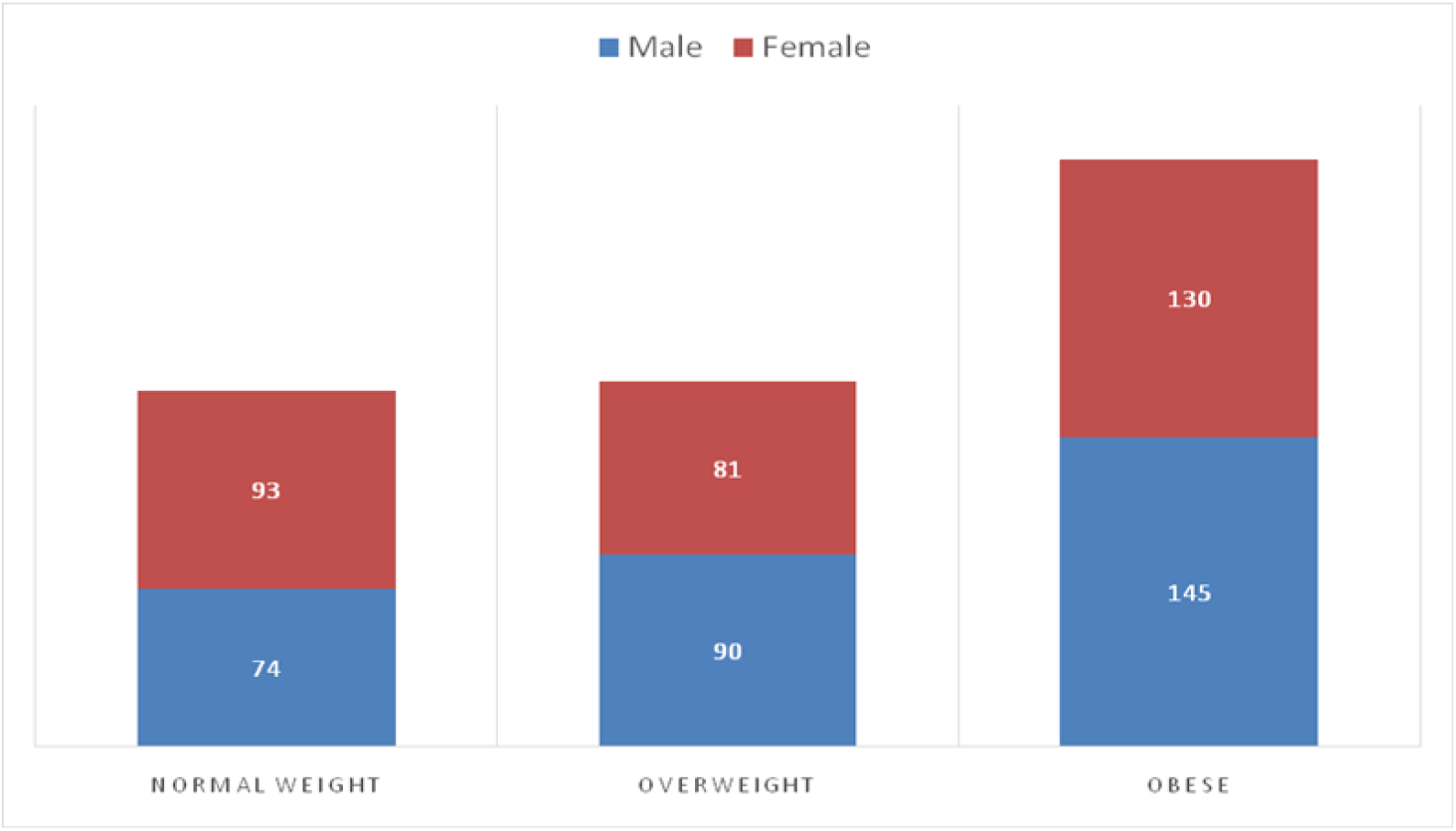
Total population 613 among them 167 normal weight,171 over weight, 275 obese.

**Table 1:**
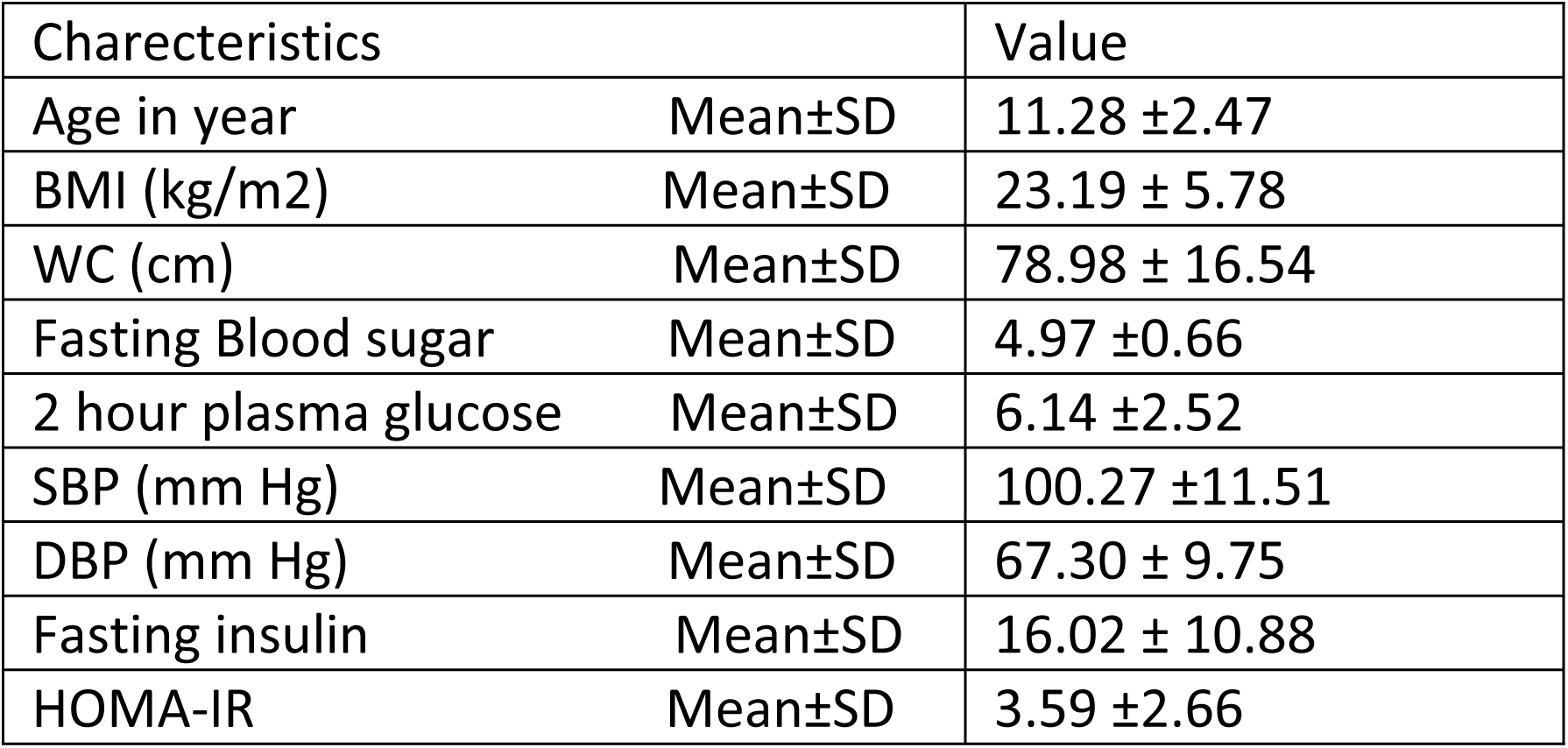
Demographic characteristics of study population.

### HOMA-IR threshold above 85^th^ percentile for normal weight children

The cut off value of HOMA-IR determined by percentile criteria in normal weight children according to grnder and puberty without related components of metabolic syndrome given in Table2. HOMA –IR cut- off values were 2.4 in total population,2.6 in girl,2.3 in boys, 2.3 in pre pubertal,2.6 at pubertal period at 85^th^ percentile. We found no significant gender, pubertal difference in HOMA-IR threshold at 85^th^ percentile.

**TABLE 2:**
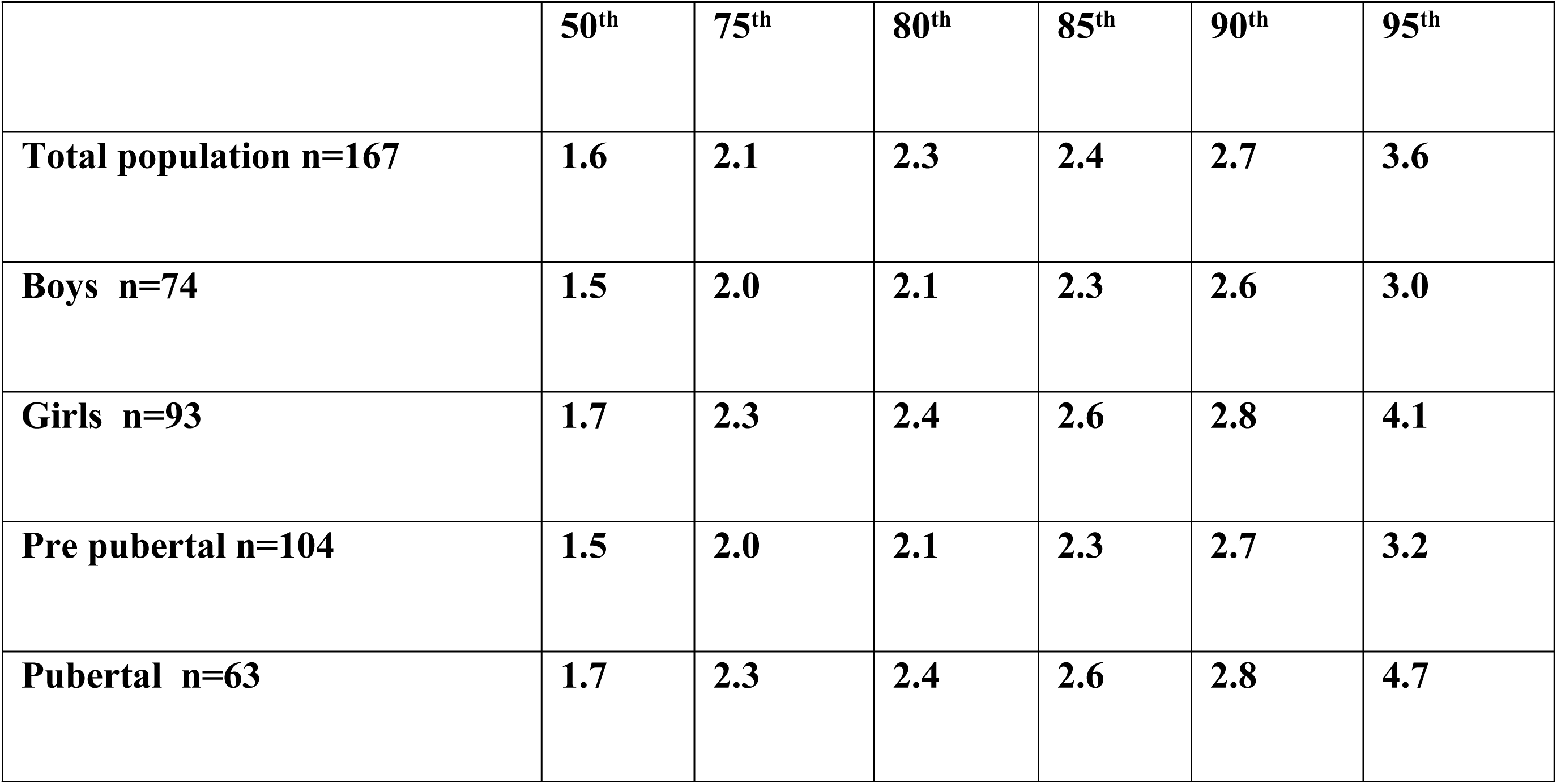
Cut off value of HOMA-IR determined by percentile criteria in normal weight children according to grnder and puberty without related components of metabolic syndrome.

### The cut-off value of HOMA-IR determined by ROC curve analysis

Among subjects with metabolic syndrome diagnosed by the modified ATP III definition cut off value for the total participants was 2.16 (sensitivity=91.8%, specificity=60.5%, Area under the curve 0.842) (fig 3). When considering sex and puberty the cut off 2.2 for boys(sensitivity=91.7%, specificity=60.0%, Area under the curve 0.843), 2.13 for girls (sensitivity=91.9%, specificity=60.0%, Area under the curve 0.838), 2.0 for pre pubertal period (sensitivity=93.9%, specificity=62.1%, Area under the curve 0.865), 2.4 for pubertal period(sensitivity=93.2%, specificity=60.1%, Area under the curve 0.842)(fig 2,3,4,5,6 respectively).

**Fig 2:**
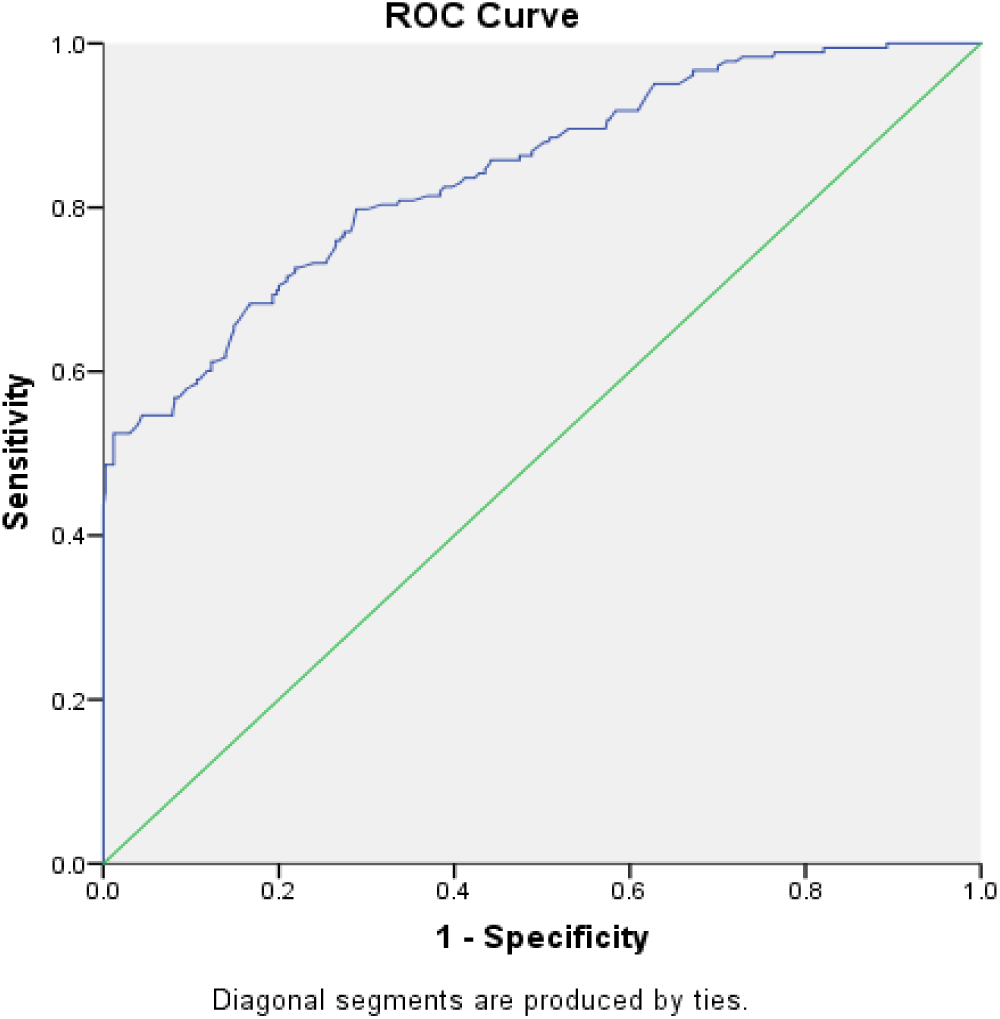
ROC curve analysis of HOMA-IR determined by metabolic syndrome ATP III criteria in Bangladeshi children. ROC curve shows optimal cut off value of HOMA-IR in Bangladeshi children to be 2.16 with area under curve 0.842 and p value <0.0001

**Fig 3:**
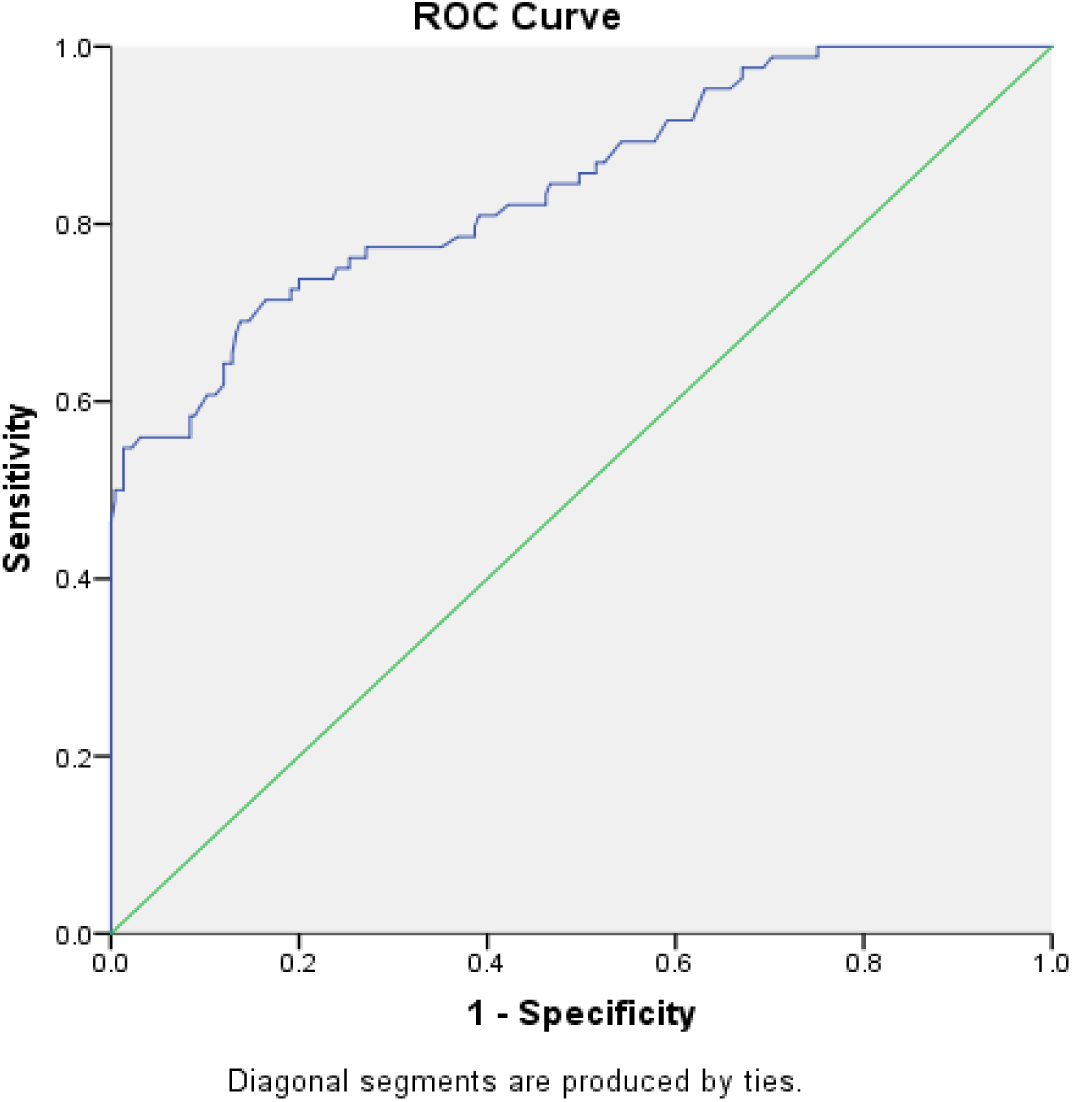
ROC curve analysis of HOMA-IR determined by metabolic syndrome criteria in boys. ROC curve shows optimal cut–off value of HOMA–IR in boys to be 2.2 with area under curve 0.843 and p-value <0.0001.

**Fig 4:**
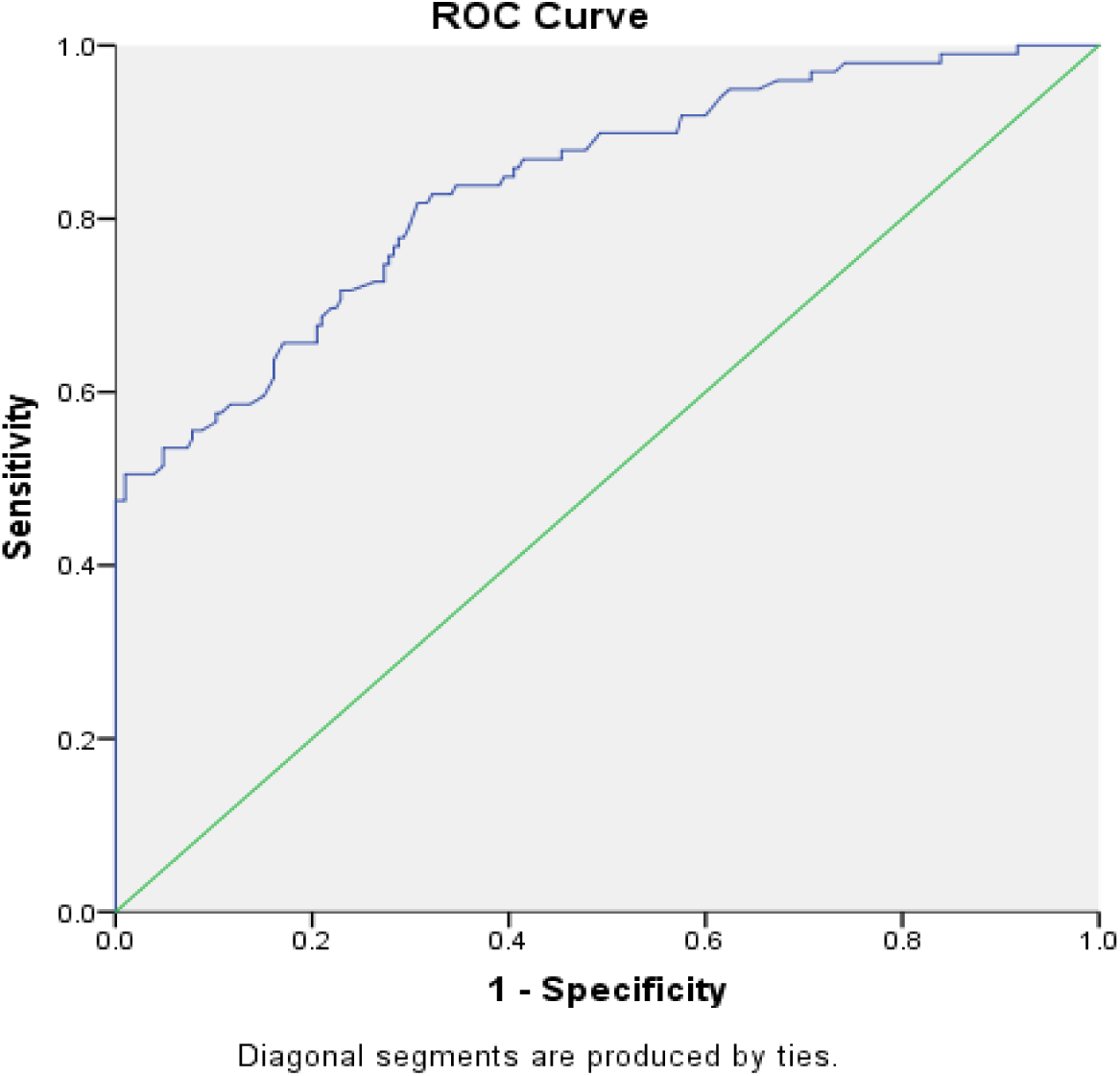
ROC curve analysis of HOMA-IR determined by metabolic syndrome ATP III criteria in girls. ROC curve shows optimal cut off value of HOMA-IR in girls to be 2.13 with area under curve 0.838 and p-value <0.0001

**Fig 5:**
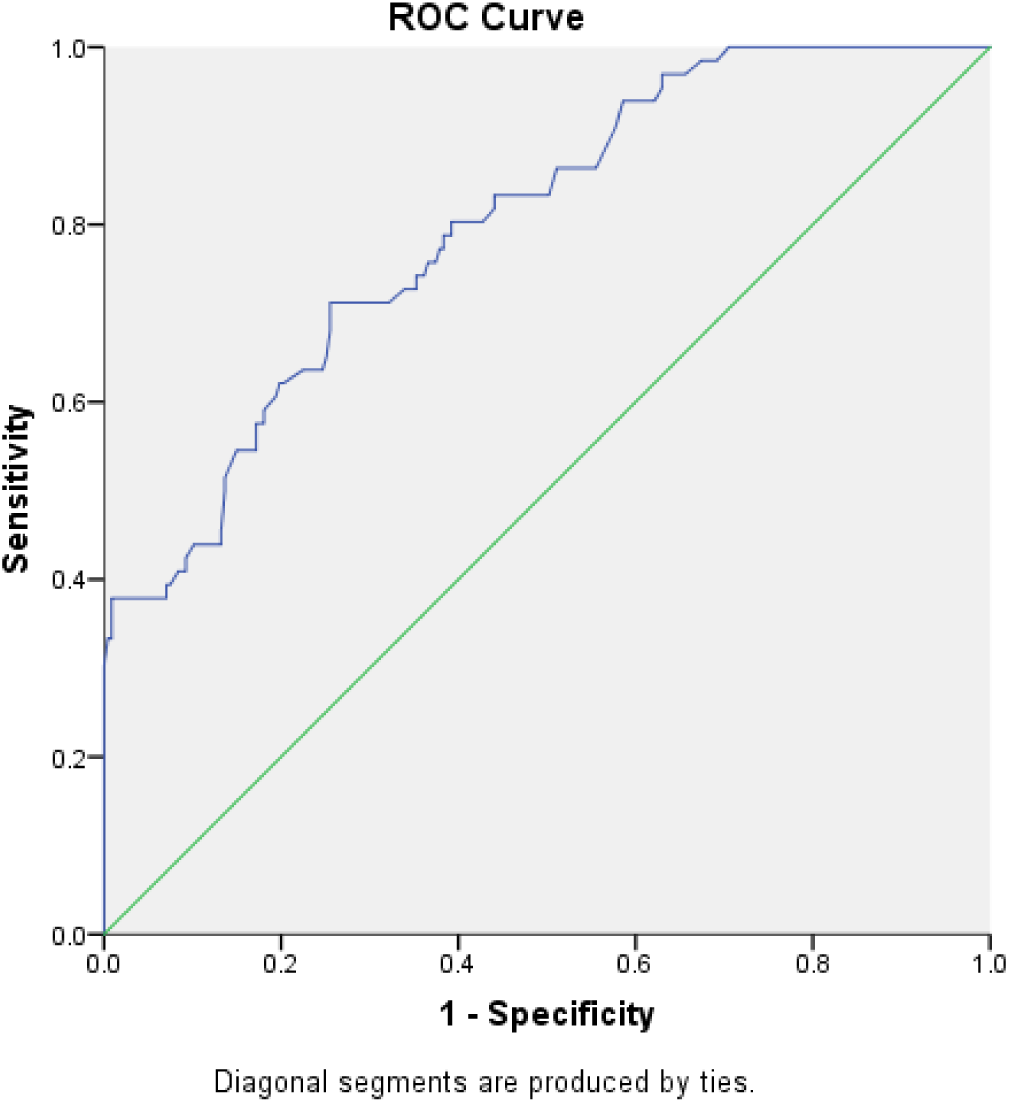
ROC curve analysis of HOMA-IR determined by metabolic syndrome ATP III criteria in pre-pubertal children. ROC curve shows optimal cut off value of HOMA-IR in pre-pubertal children to be 2.0 with area under curve 0.799 and p-value <0.0001

**Fig 6:**
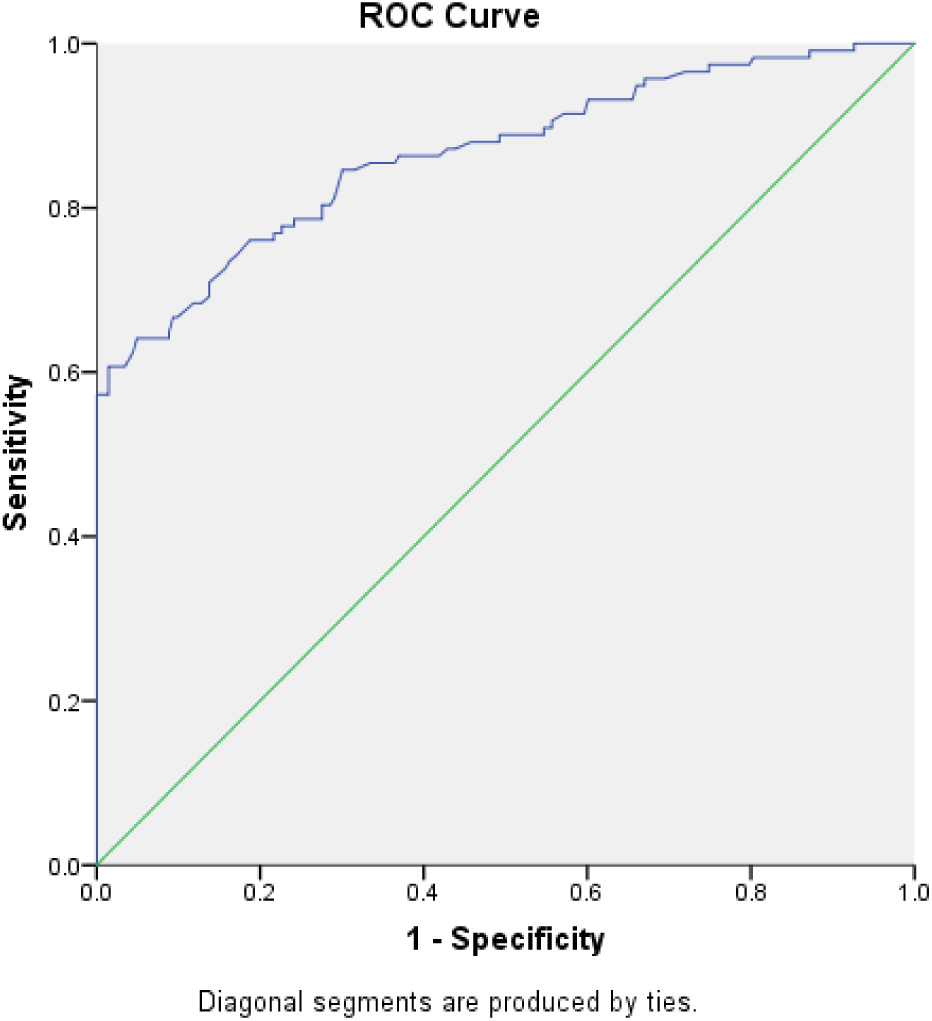
ROC curve analysis of HOMA-IR determined by metabolic syndrome criteria in pubertal children. ROC curve shows optimal cut off value of HOMA-IR in pubertal children to be 2.4 with area under curve 0.865 and p-value <0.0001

**Table 3:**
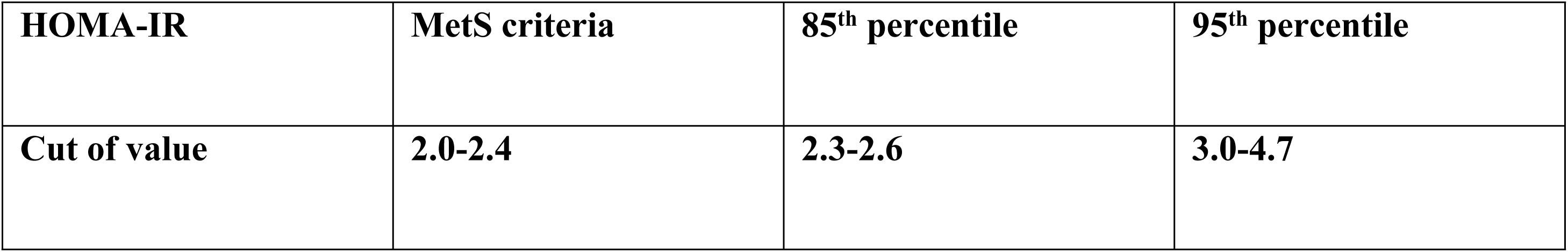
Summary cut-off value range of HOMA-IR for diagnosis of IR determined by percentile criteria and metabolic syndrome ATP III criteria irrespective of sex and puberty.

**Table 4:**
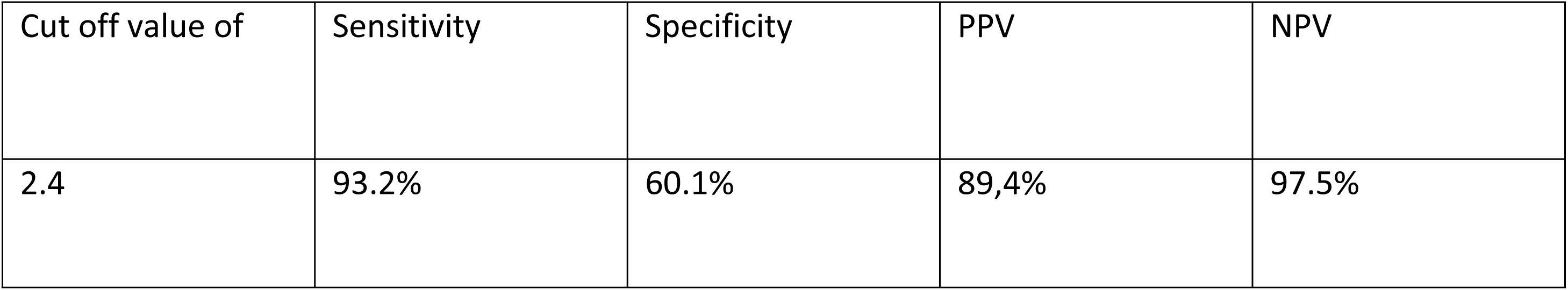
Performance HOMA-IR cut-off value 2.4 (common to metabolic syndrome criteria and P85) for diagnosis of insulin resistance in Bangladeshi children irrespective of sex and puberty.

## DISCUSSION

Insulin resistance is a key component of metabolic syndrome hence early diagnosis of insulin resistance is of great value in preventing chronic disease like type 2 diabetes mellitus, dyslipidemia, hypertension, cardiovascular disease, non-alcoholic fatty liver disease, polycystic overy syndrome in adulthood. Insulin resistance is the first sign of disturbed glucose metabolism and it revealed that appreciable β-cell dysfunction occur before glucose tolerance or fasting glucose levels become impaired (24).

In present study we used most commonly adapted method for determining insulin resistance which are obtained from practical formula, currently in our country, there is no data defining the optimal cut-off values of HOMA-IR for insulin resistance in children. So, to find out the optimal cut-off point of HOMA – IR we used percentile criteria and metabolic syndrome criteria. The International Diabetes Federation (IDF) released the first definition of MS in children and adolescents in 2007. However, it positions central obesity as an essential element, this almost rules out the possibility of diagnos ing MS in a normal weight individual (25). so we used modified ATP III criteria (Adult Treatment Panel III) of metabolic syndrome for identifying insulin resistance in both sex. The component of this criteria are Waist Circumference, fasting plasma glucose, Hypertension, low HDL, high cholesterol and high triglyceride

Rocco et al. (2011) conducted a study on 319 Brazillian children and found HOMA-IR cut-off value 2.1 at 85th percentile whereas Yin et al. (2013) conducted a study on 3203 Chinese children and found HOMA-IR cut-off value 3.0 at 95^th^ percentile, Shashajet al. (2015) conducted a study in Rome (Italy) on 2573 Caucasian children, where they found the HOMA–IR cut–off value 3.02 at 75th percentile and Tresaco et al.(2005) found HOMA-IR cut-off value 2.07 at 50^th^ percentile and 2.83 at 75th percentile. In Chinese children the cut-off value of HOMA– IR in percentile criteria dropped from 3.0 to 2.3 and in Brazillian children cut-off value dropped from 2.1 to 1.65 in girls 1.95 in boys when metabolic syndrome was taken into account (15,16).

In present study HOMA–IR cutoff value found 3.6 at 95th percentile; the sensitivity and specificity were 78.6%, and 72.9% respectively. However at 85^th^ percentile the cut–off value was 2.4 which is lower than 3.6 but had higher sensitivity and specificity 93.2% and 60.1%.

HOMA-IR values ranging from 1.65 to 3.16 with high sensitivity and specificity have been reported as cut-off for identifying IR in various studies (15,24–26) but all these studies were held on small samples of obese children and adolescents, hence are likely to report higher cut-off values. But our cut-off value derived from a homogenous mix of normal-weight, overweight, and obese children (both sex and puberty) with relatively large sample size.

According to Carrazzone et al.(2004) screening tests require high sensitivity and high NPV and whereas a diagnostic test require higher specificities and high PPV (27). Most of the study for identifying insulin resistance in children, cut-off value obtained from both sex instead of adjustment of sex and puberty. In Present study showed high sensitivity and moderate specificity at cut-off value by ROC curve, when it is determined by metabolic syndrome criteria and calculated by Youden index (max of sensitivity+specificity). In present study boys shows cut-off 2.2, AUC 0.843 with sensitivity 91.7%, NPV 96.3% and specificity 60.0%, in girls cut-off value 2.13, AUC 0.838, with sensitivity 91.9%, NPV 98.4% and specificity 60.0%, in Prepubertal child cut-off value 2.0, AUC 0.799, sensitivity 93.9%, NPV 98.4% and specificity 62.1% and Pubertal child cut-off 2.4, AUC 0.865, sensitivity 93.2%, NPV 99.5% and specificity 60.1%. Yin et al.(2013) conducted a cross sectional study on 3203 Chinese children, which is consistent with our study, where they found optimal cut-off point for diagnosis of Metabolic syndrome was 2.3 in total participants,1.7 in pre-pubertal and 2.6 in pubertal children by ROC curve which yield high sensitivity and moderate specificity.

The cut-off value of HOMA-IR at 85th percentile ranges between 2.3 to 2.6 and cut-off by metabolic syndrome criteria ranges between 2.0 to 2.4 ; therefore we suggest 2.4 as the cut–off value of insulin resistance, as because it is supported by both the percentile criteria and metabolic syndrome criteria where sensitivity and NPV found high almost similar. Early detection of insulin resistance will help to raise awareness, diagnose early and manage effectively to prevent the cardio metabolic complications and type 2 diabetes mellitus among children.

## Conclusion

This study suggests 2.4 is the optimal cut-off value of insulin resistance because it supported by both the percentile citeria and metabolic syndrome criteria.

Previously insulin resistance was exclusive to adults, however studies indicate that insulin resistance also present in children (11,16–18).

## Data Availability

All relevant data are within the manuscript and its Supporting Information files.

